# Overall and sex-specific effect of berberine for dyslipidemia: systematic review and meta-analysis of placebo-controlled trials

**DOI:** 10.1101/2022.06.20.22276676

**Authors:** Joseph E Blais, Xin Huang, Jie V Zhao

## Abstract

**Background:** Berberine is a nutraceutical that may improve lipid profiles. Berberine may also affect sex hormones and exert sex-specific effects, which has been overlooked.

**Objectives:** To comprehensively review the efficacy and safety of lipid-lowering effect of berberine with consideration of potential sex disparity.

**Methods:** Eligible studies were randomized controlled trials in adults that compared berberine versus placebo and measured blood lipids or lipoproteins. Studies were identified from Medline, Embase, Wanfang, CNKI, two clinical trial registries and previous systematic reviews. Mean differences (MD) were estimated using inverse variance weighting with random effects models. Risk of bias was assessed using the Cochrane risk of bias tool for randomized trials.

**Results:** 16 studies were included with treatment of 4 to 24 weeks. Berberine reduced low-density lipoprotein (LDL) cholesterol (−0.45 mmol/L, 95% CI -0.60 to -0.31, 12 studies, n=1,224), total cholesterol (−0.47 mmol/L, 95% CI -0.61 to -0.33, 15 studies, n=1,397), triglycerides (−0.32 mmol/L, 95% CI -0.44 to -0.19, 16 studies, n=1,421) and apolipoprotein B (−0.25 mg/dL, 95% CI -0.40 to -0.11, 2 studies, n=127). Berberine increased high-density lipoprotein (HDL) cholesterol by 0.06 mmol/L (95% CI 0.00 to 0.12, 13 studies, n=1,248). Notably, the effect on HDL cholesterol was different in women (0.11 mmol/L, 95% CI 0.09 to 0.13) from that in men (−0.07 mmol/L, 95% CI -0.16 to 0.02). Gastrointestinal adverse events were the most frequently reported adverse events.

**Conclusions:** Berberine decreased LDL cholesterol, triglycerides, and apolipoprotein B, with a potential sex-specific effect on HDL cholesterol. Large-scale trials considering sex disparity are required.

## Introduction

Dyslipidemia is highly prevalent. Elevated non-HDL cholesterol was attributable for 3.9 million deaths from ischemic heart disease and stroke in 2017.(1) Globally, statins are the most common treatment for dyslipidemia.(2) However, nearly 10% of patients treated with statins experience statin intolerance and the odds of statin intolerance are greater in women and Asians.(3) Clinicians need to consider patient preferences in their decision making, and some patients may decide to use a non-prescription nutraceutical to manage their dyslipidemia.

Berberine is a pharmacologically active plant alkaloid that is present in several plants used in herbal medicine: goldenseal (*Hydrastis canadensis*), Coptis or goldenthread (*Coptis chinensis*), Oregon grape (*Berberis aquifolium*), barberry (*Berberis vulgaris*), and tree turmeric (*Berberis aristata*).(4) Nutraceutical supplements of berberine, produced using chemical synthesis, are widely available in Europe, North America, and Asia through local and online retailers. Berberine has been recommended as a complement to statins by both the International Lipid Expert Panel and the 2019 European Atherosclerosis Society/European Society of Cardiology Guidelines for the treatment of dyslipidemia in statin-intolerant patients,(5 24, 6) yet, these guidelines have not provided explicit recommendations on the use of berberine because of the lack of high-quality evidence.

Prior reviews of berberine have several limitations as they included studies that did not make comparisons to placebo, compared berberine to diverse controls (e.g., statins, oral hypoglycemics, lifestyle modification), assessed extracts from the barberry plant or defined the intervention as combination nutraceutical products containing berberine such as Armolipid Plus®.(7-13) Moreover, the bioavailability of the different berberine preparations is a matter of debate.(14) Considering this, it is crucial to update and synthesize similar trials that assess the efficacy of purified berberine, the formulation most often used as a nutraceutical or supplement.

Notably, it is increasingly realized that lipid-lowering drugs, such as statins, may exert sex-specific effects, possibly through sex hormones.(15) Berberine may also affect sex hormones, and therefore exert sex-specific effects. In our randomized controlled trial in Chinese men, berberine increased serum testosterone, whereas trials in women with polycystic ovary syndrome (PCOS) showed berberine decreased testosterone concentrations.(16-18) To our knowledge, the sex-specific effect has not been considered in previous systematic reviews of berberine.

The objectives of this systematic review were to assess whether oral berberine monotherapy reduces lipids and lipoproteins compared with placebo in adults and to examine the potential sex-specific efficacy of berberine. We also assessed the safety of berberine. This review can inform patients and clinicians of the totality of the currently best-available evidence for berberine, it can meet the needs of evidence-based clinical practice guideline recommendations for use of berberine for treatment of dyslipidemia,(19) and it can inform future research on berberine as an intervention for dyslipidemia and preventing atherosclerotic cardiovascular events.

## Methods

### Review methods and registration

We did a study-level systematic review and meta-analysis of randomized controlled trials. The study protocol was as registered in PROSPERO (<u>CRD42021293218</u>) prior to completing the initial literature search.(20) This report adheres to the Preferred Reporting Items for Systematic reviews and Meta-Analyses (PRISMA) 2020 statement.(21) Covidence systematic review software (Veritas Health Innovation, Melbourne, Australia) was used to manage the entire review process and deduplicate studies.

### Eligibility criteria

All randomized controlled trials, regardless of publication language, that compared berberine with placebo in participants ≥ 18 years and measured a lipid or lipoprotein outcome were eligible for inclusion. We excluded studies that 1) did not report any lipid or lipoprotein outcome measure; or 2) that investigated plant extracts containing berberine, since we wanted to evaluate the effect of synthetic berberine that is commercially formulated as tablets and capsules; or 3) that did not compare berberine to placebo (e.g., no drug, lifestyle intervention); or 4) that assessed berberine in combination with other nutraceutical compounds or lipid-modifying drugs; or 5) that were observational studies, unpublished reports, conference abstracts, reviews, animal or cell studies.

### Information sources and search strategy

We searched four databases, MEDLINE via Ovid (1946 to December 10, 2021), Embase via Ovid (1947 to December 10, 2021), Wanfang (inception until December 15, 2021), and CNKI (China National Knowledge Infrastructure) (inception until December 15, 2021), and two clinical trial registries, ClinicalTrials.gov (inception until December 14, 2021) and the World Health Organization International Clinical Trials Registry Platform (inception until December 14, 2021). These searches were supplemented by reviewing the reference lists of previous systematic reviews.(7-11, 13) Search terms included Participant Intervention Comparator Outcomes (PICO) terms such as berberine, dyslipidemia, hyperlipidemia, low-density lipoprotein (LDL) cholesterol, triglycerides (TG), total cholesterol (TC), high-density lipoprotein (HDL) cholesterol, and apolipoprotein B (apoB). The detailed search strategy and results are included in the Supplementary Material File 1.

### Study selection process

Two reviewers (JEB and XH) independently screened the title and abstract of each study and discrepancies were independently assessed by a third reviewer (JVZ). After screening, two reviewers independently reviewed the full text of each study and determined eligibility for inclusion. If excluded, the reviewers also specified the primary reason for study exclusion, and discussed to reach a consensus on the primary reason for study exclusion (Supplementary Material File 2). Consensus for inclusion and exclusion reasons was also achieved through discussion with a third reviewer.

### Data collection process and data items

We developed a data extraction template within Covidence and piloted the extraction process with one study (see Supplementary Material File 3). One reviewer independently extracted the data from each record and a second reviewer independently checked the extraction data. Discrepancies were resolved through discussion within the two reviewers.

The primary objective of the study was to assess the efficacy of berberine in lowering lipids and lipoproteins. The outcomes we used were the differences between berberine and placebo at 12 weeks after randomization in TC, LDL cholesterol, non-HDL cholesterol, HDL cholesterol, TG, and apoB. If outcomes were not measured at 12 weeks, we used the last measurement within each study. Data items that were extracted for each study included author contact information; year of study publication; country of study; funding source; characteristics of the intervention; characteristics of the placebo; treatment duration, eligibility criteria; baseline participant characteristics; outcome measurement values, units, and time points. For trials with more than two treatment arms, data were extracted for the placebo and berberine groups.

The secondary objective of the study was to assess the safety of berberine, where we recorded any reported adverse events. In our pilot search, we identified commonly reported adverse events reported in previous reviews comparing berberine to other interventions such as statins, including gastrointestinal (e.g., nausea, diarrhea, abdominal discomfort) and muscle-related adverse events (e.g., muscle pain, myopathy). We listed these events in the data extraction form and put down the number of these events and number of participants with each adverse event (when reported) in our systematic review. The items we extracted included the type of adverse events, number of adverse events and number of participants experiencing each adverse event as reported in the original trials.

### Study risk of bias assessment

Two reviewers assessed the risk of bias for each study using the Cochrane RoB 2 tool.(22) Risk of bias was judged as ‘low risk of bias’, ‘some concerns’, or ‘high risk’ of bias across five distinct study domains (bias arising from the randomization process, bias due to deviations from the intended interventions, bias due to missing outcome data, bias in measurement of the outcome, and bias in selection of the reported result) and for the overall study risk of bias.

### Synthesis methods

We pooled results for each outcome if there were at least two similar studies that assessed it. We calculated the changes in each of the berberine and placebo groups at baseline and post-treatment and compared the difference in changes between the two groups. We extracted the mean, SD for baseline and post-treatment from original articles and calculated the mean and SD of the change from baseline according to the following equation from the Cochrane handbook:

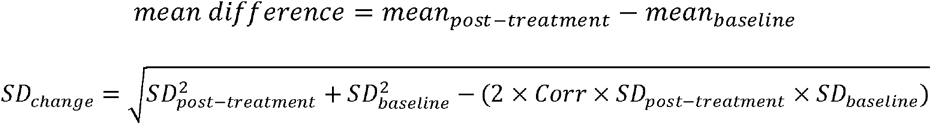

The correlation coefficients (Corr) for TC, LDL, HDL, and TG were calculated from Zhang et al. as this study reported means and SD for change as well as for baseline and post-treatment. (23) Corr for apoB was assumed to be 0.5

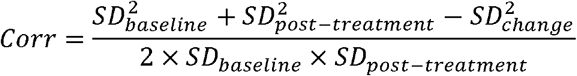

For studies that provided detailed results, we extracted their mean difference and SD difference directly.(23, 24) For studies that reported medians and interquartile ranges (IQR), we estimated the mean and SD as described in Luo and Wan.(25, 26)

Study level estimates for lipid outcomes were pooled as mean differences (MD) and 95% confidence intervals (CI) using a random effects model with inverse-variance method. If multiple analyses were reported, we used the per-protocol estimate for meta-analysis. We used the R package *meta* (version 5.2-0) and the “metacont” function to estimate MDs and the “forest” function to generate forest plots. Between study heterogeneity was assessed visually and statistically using I^2^. Funnel plots and Egger’s test were used to assess potential publication bias. We estimated the overall effect by using the trim and fill method to add back the “missing” studies using the “trimfill” function.

To investigate the potential difference by sex, we also conducted the analysis in studies only in men, only in women, or in both men and women, and compared the estimates of meta-analysis using Q-test based on analysis of variance. We also conducted additional sensitivity analyses to examine if there is any heterogeneity according to trial duration (≤ 12 weeks, > 12 weeks), berberine dose (<1000 mg/day, ≥ 1000 mg/day) and ethnicity (Asian, non-Asian). All the analyses were conducted using R (R Foundation for Statistical Computing, Vienna, Austria; Version 1.4.1717).

### Certainty assessment

The quality of the evidence was evaluated and reported using the Grading of Recommendations Assessment, Development and Evaluation (GRADE) working group methodology.(27) Two reviewers assessed the following factors for each outcome: risk of bias, imprecision, inconsistency, indirectness, and publication bias. GRADEpro GDT software was used to create a summary of findings table for the key primary and secondary outcomes.(28)

## Results

### Study selection

After removing duplicate records and screening abstracts, 54 studies were assessed in full text (Figure 1). The most common reason for excluding studies was that they assessed the wrong intervention such as extracts of barberry (that contain berberine), or did not compare berberine to placebo. We identified two publications from the same authors that we considered to be the same study, with one study published in Chinese and the other in English.(29, 30) To avoid duplication, we only present data from the earlier publication as the sample size was larger and the study report was more detailed.

**Figure 1:**
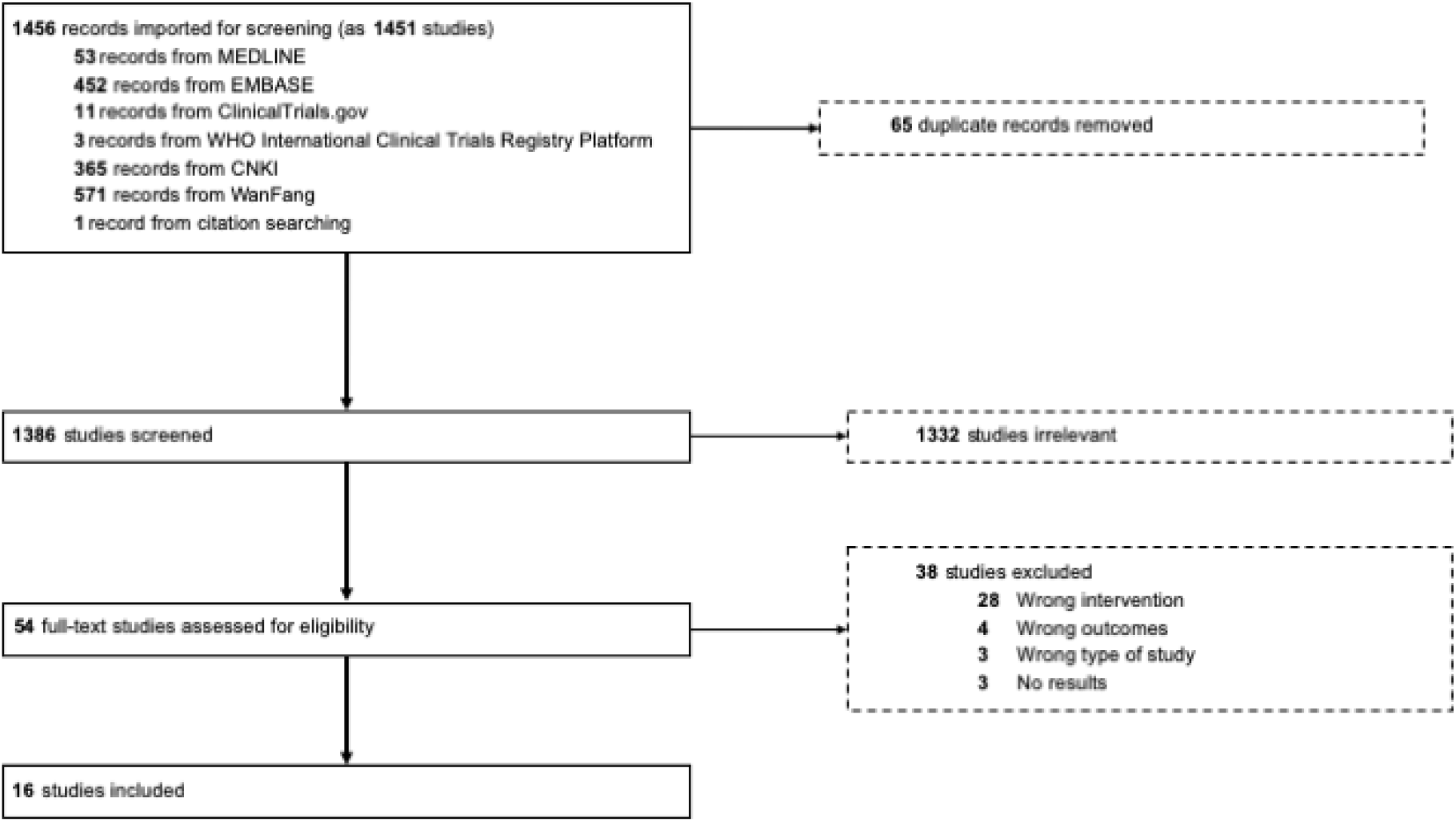
Preferred Reporting Items for Systematic reviews and Meta-Analyses (PRISMA) flow chart of study inclusion in the systematic review and meta-analysis

### Study characteristics

A total of 16 studies met the eligibility criteria with 1,529 participants (n= 761 berberine; 768 = placebo; Table 1; Supplementary file 3 p. 2 shows records for each study).(16-18, 23, 24, 29, 31-40) All studies were conducted in young or middle-aged adults and none of the studies enrolled participants with a history of coronary artery disease or stroke. Thirteen studies (81%) were conducted in mainland China and Hong Kong. Trial duration ranged from 4 to 24 weeks. Patients enrolled in the studies most frequently had type 2 diabetes mellitus,(23, 24, 32, 35, 39, 40) PCOS,(17, 18, 33, 38) dyslipidemia,(16, 31, 34, 36) or schizophrenia.(29, 37) One study exclusively enrolled men,(16) four studies exclusively enrolled women with PCOS,(17, 18, 33, 38) and the remaining studies enrolled both men and women.

### Risk of bias assessment

We considered the risk of bias as “low” for 12 studies, “some concerns” for three studies and “high” for one study. The risk of bias assessment for each domain and at the overall study-level are included in Supplementary Table 2.

### Findings on efficacy

A summary of the key results and certainty of evidence is presented in Table 2. The mean difference for berberine versus placebo was -0.47 mmol/L (95% CI -0.61 to -0.33, I^2^=84%; 15 studies, n=1,397 participants; moderate certainty; Figure 2A) for TC, -0.45 mmol/L (95% CI - 0.60 to -0.31, I^2^=80%; 12 studies, n=1,224 participants; moderate certainty; Figure 2B) for LDL cholesterol, -0.32 mmol/L (95% CI -0.44 to -0.19, I^2^=79%; 16 studies, n=1,421 participants; moderate certainty; Figure 2C) for TG, 0.06 mmol/L (95% CI 0.00 to 0.12, I^2^=89%; 13 studies, n=1,248 participants; low certainty; Figure 2D) for HDL cholesterol, -0.25 mmol/L (95% CI -0.40 to -0.11, I^2^=82%; 2 studies, n=127 participants; low certainty; Figure 2E) for apoB.

**Figure 2:**
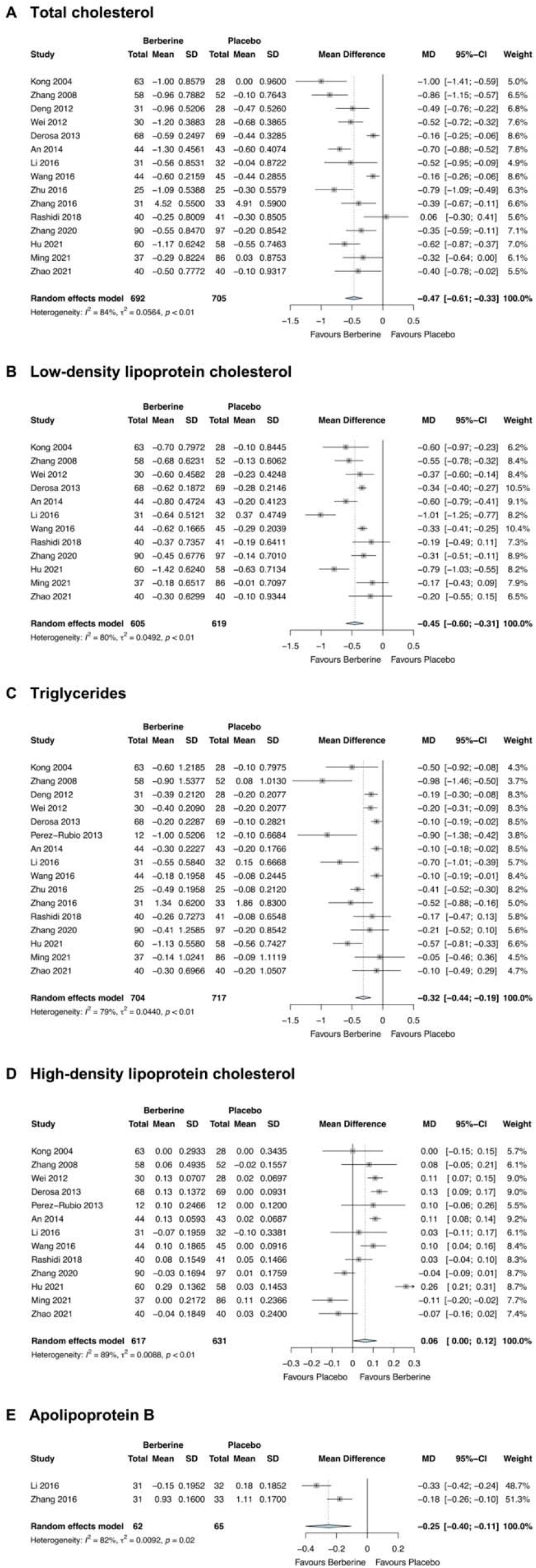
Forest plots for total cholesterol (A), low-density lipoprotein cholesterol (B), triglycerides (C) high-density lipoprotein cholesterol (D), and apolipoprotein B (E).

Although there was considerable statistical heterogeneity for each outcome, the direction and magnitude of the effect of berberine was consistent for LDL cholesterol and apoB. Greater heterogeneity in the direction of effect was visible for HDL cholesterol, while the magnitude of TG reduction varied among studies.

Funnel plots showed fewer trials on the right-hand side where berberine increased TC and TG (Supplementary Figure 1A and 1C), and Egger’s test provided significant evidence of asymmetry (P=0.0109 for TC; P=0.0106 for TG). Trim and fill revised the mean difference to -0.23 mmol/L (95% CI -0.42 to -0.05) for TC and -0.20 mmol/L (95% CI -0.37 to -0.03) for TG. There was limited evidence of asymmetry for LDL and HDL cholesterol (Supplementary Figure 1B and 1D). The number of included studies that reported apoB was insufficient to generate a funnel plot.

### Findings on safety

Adverse event reporting in the eligible studies was inconsistent and heterogeneous and precluded meta-analysis (see Supplementary File 5 for extracted adverse event data). Two studies did not report adverse events.(17, 40). The frequency of participant withdrawal from the study or discontinuation of the treatment due to adverse events ranged from 2 to 6% of allocated patients in four studies.(18, 24, 33, 37) Although the overall number of any adverse events or proportion of patients with an adverse event were similar in five studies (23, 24, 29, 35, 37), gastrointestinal adverse events, such as constipation, diarrhea, and nausea, appeared to occur more frequently in participants randomized to berberine (very low to low certainty).(16, 18, 23, 29, 31, 32) No serious adverse events were reported in participants randomized to berberine. There were also no reports of muscle pain in any of the eligible studies. Derosa et al reported no cases of myopathy and no increases in creatine phosphokinase.(34) Two cases of liver function abnormalities were reported in one study in the berberine group,(29) although two studies reported no increase or a decrease in liver transaminases with berberine.(32, 34) Given its ability to reduce blood glucose, berberine was associated with greater hypoglycemic events and fewer hyperglycemic events than placebo in 4 studies.(24, 29, 37, 39)

### Subgroup Analyses

Stratified analyses suggested sex-specific differences in the effect of berberine on HDL cholesterol (women: 0.11 mmol/L, 95% CI 0.09 to 0.13; men: -0.07 mmol/L, 95% CI -0.16 to 0.02; women and men: 0.06 mmol/L, 95%CI -0.01 to 0.13; P <0.01; Figure 3D). There was less evidence of sex-specific differences for TC (P=0.16), LDL cholesterol (P=0.34) and TG (P= 0.21), although heterogeneity was greatly lowered in the sex-specific analysis, where the heterogeneity was lowest in the studies that enrolled women with PCOS for the outcomes of TC and LDL cholesterol (Figure 3A and 3B).

**Figure 3:**
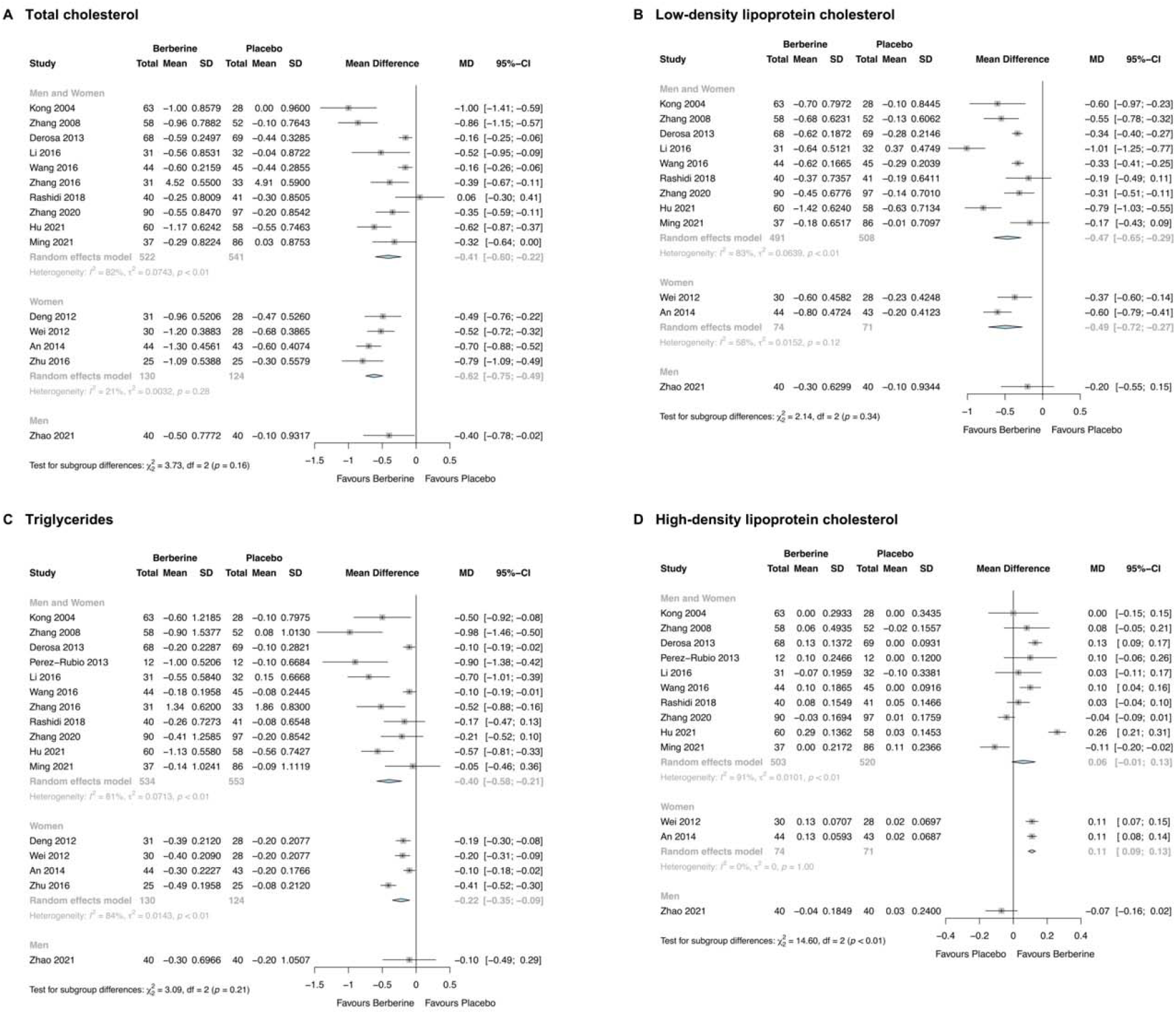
Forest plot stratified by sex for total cholesterol (A), low-density lipoprotein cholesterol (B), triglycerides (C) and high-density lipoprotein cholesterol (D).

There was no evidence of a difference of the effect of berberine on TC (P=0.82), LDL (P=0.09), TG (P=0.56), and HDL cholesterol (P=0.47) at different treatment durations (≥ 12 weeks, <12 weeks; Figure 4). There was no evidence of a difference in the effect of berberine on TC (P=0.80), LDL cholesterol (P=0.48), TG (P=0.75) and HDL cholesterol (P=0.49) at different treatment doses (<1000 mg/day, ≥1000 mg/day; Supplementary Figure 2). There was evidence of potential ethnic differences in the therapeutic effects of berberine on TC (Asian: -0.50 mmol/L, 95%CI -0.64 to -0.36; non-Asian: -0.16 mmol/L, 95%CI -0.25 to -0.06; P <0.01; Supplementary Figure 3) and on HDL cholesterol (Asian: 0.05 mmol/L, 95%CI -0.01 to 0.11; non-Asian: 0.13 mmol/L, 95%CI 0.09 to 0.17; P=0.04). No significant ethnic differences were observed for LDL cholesterol (P=0.13) or TG (P=0.70) (Supplementary Figure 3).

**Figure 4:**
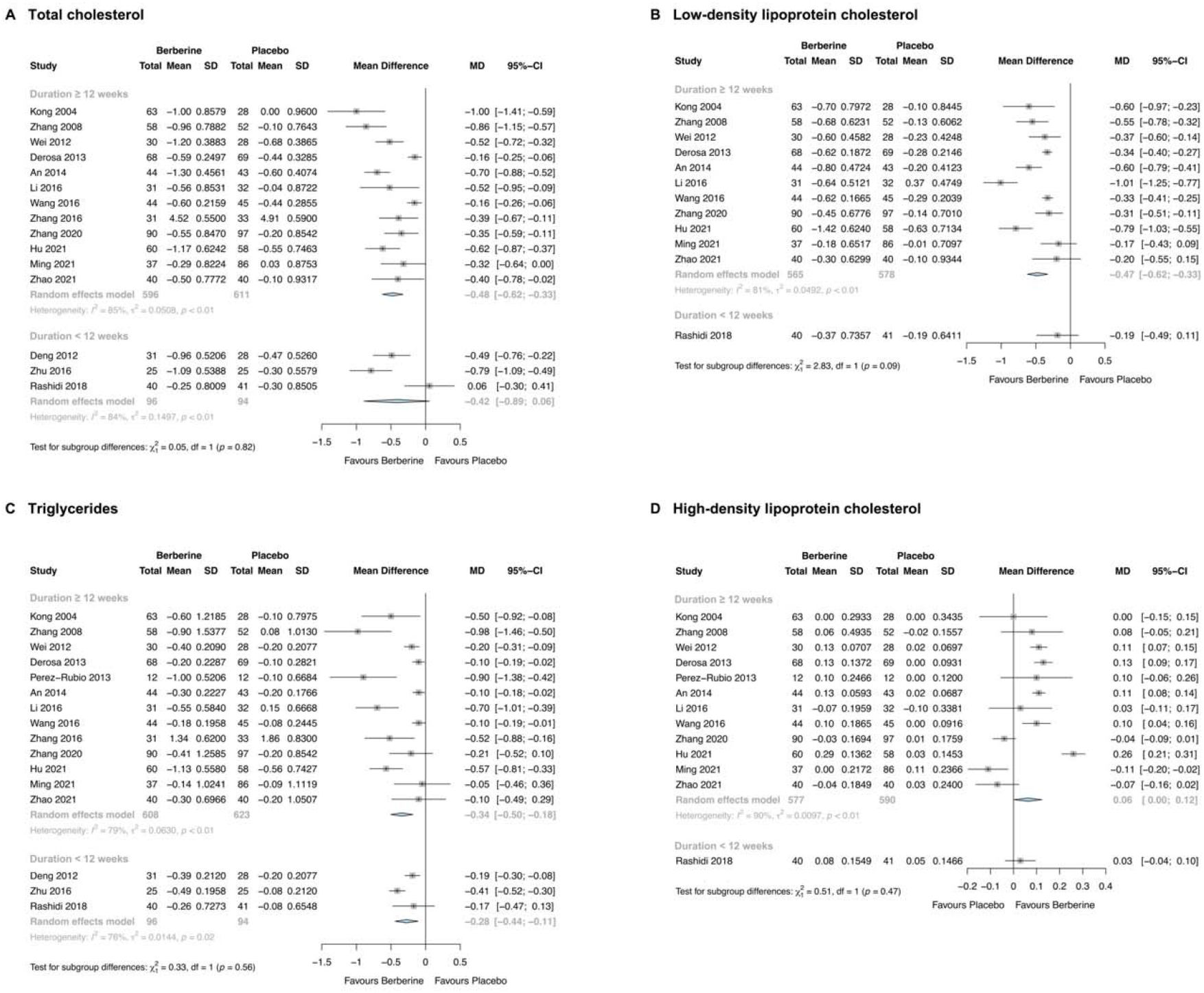
Forest plot stratified by study duration for total cholesterol (A), low-density lipoprotein cholesterol (B), triglycerides (C), and high-density lipoprotein cholesterol (D).

## Discussion

### Interpretation of the results

This systematic review and meta-analysis incorporated evidence comparing purified berberine and placebo demonstrates a short-term effect of berberine for improving lipids and lipoproteins in young and middle-aged adults from diverse populations without established cardiovascular disease. Our findings are consistent with previous systematic reviews and meta-analyses that included plant extracts of berberine as the intervention, which partly adds to the debate by showing that different preparations of berberine may all exert lipid-lowering effects. Using placebo as comparison also supports the efficacy of berberine in lipid lowering as a stand-alone therapy. The mechanisms might be due to berberine enhancing the hepatic uptake of LDL particles by inhibiting proprotein convertase subtilisin/kexin type 9 (PCSK9), decreasing intestinal cholesterol absorption, increasing fecal cholesterol excretion, and enhancing hepatic cholesterol turnover.(5)

Interestingly, our study also found a sex-specific effect of berberine on HDL cholesterol, where the effect on HDL cholesterol is more pronounced in women than in men. One possible explanation is that we identified more studies in women than in men, therefore the power is larger in women than in men. It might be also explained by the effect of berberine on testosterone. As shown in our previous randomized controlled trial, berberine increases testosterone in men.(16) Furthermore, our previous Mendelian randomization study(41) and a systematic review and meta-analysis of testosterone clinical trials,(42) indicates that testosterone lowers HDL cholesterol in men, which may partly explain why we did not find an increase in HDL cholesterol in men in this systematic review and meta-analysis.

Consistent with previous systematic reviews, we did not identify any serious adverse events associated with berberine.(8, 10) Thus berberine appeared to be safe and well tolerated by trial participants when taken for up to 24 weeks. The safety of berberine is also supported by its long-history and origin: extracts of medicinal plants containing berberine have been used in ancient China for thousands of years for metabolic syndrome.

### Strengths and limitations

We searched for both published and unpublished reports in English and Chinese. Given the strong interest in berberine in China, our search strategy has likely minimized publication and language bias. Where feasible, key study outcomes were pooled and presented as absolute changes with berberine compared with placebo group. Our assessment of the certainty (quality) of the evidence will facilitate interpretation and application of the results by clinicians and patients who want to understand in what patient groups berberine may be effective and the magnitude of change in lipids and lipoproteins that can be expected with berberine.

The eligible trials in this review have four primary limitations. First, all studies assessed surrogate outcomes for the important outcome of cardiovascular events. Second, the duration of follow-up was short so clinical trials with longer duration will be needed to assess the long-term efficacy and safety of berberine. Third, there was potential bias in the reporting of outcomes. Not all studies reported all components of the standard lipid profile and the assessment and reporting of adverse events was inconsistent amongst studies. Limitations of the review include the high amount of heterogeneity among the pooled studies, which we attempted to address using several study-level subgroup analyses.

### Implications for practice and policy

This meta-analysis has synthesized the best available evidence from placebo-controlled randomized controlled trials of berberine with the consideration of sex disparity. The results can inform clinicians and clinical practice guideline panels on the effects of berberine on lipids and lipoproteins. The evidence in this study supports a potential role for berberine as a safe and effective treatment option, which could be considered in individuals who cannot or do not want other lipid-modifying drug therapies. The evidence for berberine is also strongest for women and in Asians, two important patient populations who are at greater risk of statin intolerance. In some contexts, non-prescription berberine may also be more affordable than prescription lipid-modifying drugs.

### Recommendations for future research

Future randomized trials, however, are needed to assess the long-term effects of berberine on the clinical outcomes of cardiovascular events. Trials should focus on populations where berberine has the most evidence for benefit, such as young and middle-aged women with PCOS, individuals with insulin resistance, or diabetes, and individuals who may not qualify or cannot tolerate statin therapy. Future studies should also comprehensively assess and report adverse events so that informed decisions can be made about the risks and benefits of berberine.

## Conclusion

Berberine is likely safe and effective for the treatment of dyslipidemia in individuals with type 2 diabetes, middle-aged adults, women with PCOS, and individuals with schizophrenia. Berberine should be considered in individuals with statin intolerance or for individuals who want to pursue treatment with a nutraceutical supplement. Future randomized trials are needed to determine the sex-specific efficacy of berberine for reducing the risk of atherosclerotic cardiovascular events.

## Data Availability

All data used in the analysis are included in the article and supplementary material. Data analysis code will be made available upon request to the corresponding author.

## Authors’ contributions

Conceptualization: Jie V Zhao and Joseph E Blais

Data curation: Jie V Zhao

Project administration: Joseph E Blais

Resources: Jie V Zhao

Supervision: Jie V Zhao

Validation: Jie V Zhao

Visualization: Xin Huang

Formal analysis: Xin Huang and Jie V Zhao

Writing – original draft: Joseph E Blais and Xin Huang

Writing – review & editing: All authors

## List of tables

**Table 1:** Summary of eligible studies comparing berberine and placebo

**Table 2:** Summary of findings table for berberine versus placebo for dyslipidemia

## Supplementary Material

- Supplementary File 1 - Detailed literature search (xlsx format)
- Supplementary File 2 (word format)
  - Supplementary Table 1: Exclusion criteria codes
  - Supplementary Figure 1: Trim and fill funnel plots for total cholesterol (A), LDL cholesterol (B), HDL cholesterol (C) and triglycerides (D)
  - Supplementary Figures: Additional forest plots not included in main text
    - Supplementary Figure 2: Forest plot stratified by berberine daily dose for total cholesterol (A), low-density lipoprotein cholesterol (B), triglycerides (C) and high-density lipoprotein cholesterol (D)
    - Supplementary Figure 3: Forest plot stratified by ethnicity for total cholesterol (A), low-density lipoprotein cholesterol (B), triglycerides (C) and high-density lipoprotein cholesterol (D)
- Supplementary Table 2: Study-level risk of bias assessment
- Supplementary File 3 - Data extraction template (pdf format)
- Supplementary File 4 - Adverse events raw data extraction
- PRISMA 2020 abstract checklist (for review only)
- PRISMA 2020 paper checklist (for review only)

