## Supplementary File 2 for "Overall and sex-specific effect of berberine for dyslipidemia: systematic review and meta-analysis of placebo-controlled trials"

### **Supplementary Material**

### Exclusion criteria codes

Each full-text article was assessed in priority order using the following exclusion criteria beginning at the start of the list.

| **Table 1.** Study exclusion criteria coding and examples | | |
| --- | --- | --- |
| Code | Description | Examples |
| Code 1 | Wrong type of study | - Observational studies, such as cohort studies, case reports, or case-series - Not randomized - Not controlled |
| Code 2 | Wrong intervention | - Berberine was given in combination therapy with other agents with no berberine placebo - Berberine directly compared with statin or other lipid-modifying drug - Intervention is a nutraceutical combination containing berberine as one of several ingredients - Barberry plant extract or juice - Single treatment arm |
| Code 3 | Wrong outcomes | - Did not report any lipid or lipoprotein outcome of interest |
| Code 4 | Wrong population | - Participants <18 years old |
| Code 5 | Animal studies | - In-vivo study in animals or laboratory studies in cells or tissues |
| Code 6 | Wrong study design for key questions | - Pharmacoeconomic studies - Pharmacokinetic studies (e.g., bioavailability) - Drug-drug/food interaction |
| Code 7 | Using original studies instead | - The article is a systematic review (and meta-analysis) compiled of individual study data - Conference abstract with corresponding study with full publication identified - There is another report with outcome of interest is included (e.g. full study, clinical trial registration) |
| Code 8 | Unable to obtain full paper | - Only abstract available from database and full text not located |
| Code 9 | No results available | - Final result not yet published - Premature termination of study with no outcomes reported |

### List of records identified for each study included in the review

**Kong 2004**

1. Kong W, Wei J, Abidi P, Lin M, Inaba S, Li C, et al. Berberine is a novel cholesterol-lowering drug working through a unique mechanism distinct from statins. Nat Med. 2004;10(12):1344-51. doi: 10.1038/nm1135

**Zhang 2008**

1. Zhang Y, Li X, Zou D, et al. Treatment of type 2 diabetes and dyslipidemia with the natural plant alkaloid berberine. J Clin Endocrinol Metab 2008;93(7):2559-65. doi: 10.1210/jc.2007-2404
2. ClinicalTrials.gov [Internet]. Identifier NCT00462046. Efficacy and safety of berberine in the treatment of diabetes with dyslipidemia. 2005. Available from: [https://ClinicalTrials.gov/show/NCT00462046](https://clinicaltrials.gov/show/NCT00462046)

**Deng 2012**

1. Deng H, Wei W, Guan Y. 小檗碱治疗多囊卵巢综合征伴胰岛素抵抗的研究 [The clinical study of berberine in patients with polycystic ovarian syndrome and insulin resistance]. Tianjin Medical Journal. 2012;40(10):3. Chinese. doi: 10.3969/j.issn.0253-9896.2012.10.013

**Wei 2012**

1. Wei W, Zhao H, Wang A, et al. A clinical study on the short-term effect of berberine in comparison to metformin on the metabolic characteristics of women with polycystic ovary syndrome. Eur J Endocrinol 2012;166(1):99-105. doi: 10.1530/eje-11-0616

**Derosa 2013**

1. Derosa G, D'Angelo A, Bonaventura A, et al. Effects of berberine on lipid profile in subjects with low cardiovascular risk. Expert Opin Biol Ther 2013;13(4):475-482. doi: 10.1517/14712598.2013.776037

**Perez-Rubio 2013**

1. Pérez-Rubio KG, González-Ortiz M, Martínez-Abundis E, Robles-Cervantes JA, Espinel-Bermúdez MC. Effect of berberine administration on metabolic syndrome, insulin sensitivity, and insulin secretion. Metab Syndr Relat Disord 2013;11(5):366-369. doi: 10.1089/met.2012.0183

**An 2014**

1. An Y, Sun Z, Zhang Y, et al. The use of berberine for women with polycystic ovary syndrome undergoing IVF treatment. Clin Endocrinol (Oxf) 2014;80(3):425-431. doi: 10.1111/cen.12294
2. An Y, Zhang Y, Lyu H, Li L, Sun Z. 小檗碱对行体外受精-胚胎移植的多囊卵巢综合征患者临床,内分泌,代谢指标及妊娠结局的影响 [Effect of berberine on clinical, metabolic and endocrine index and pregnancy outcome in women with polycystic ovary syndrome undergoing IVF treatment]. Modern Journal of Integrated Traditional Chinese and Western Medicine 2016;25(5):5. Chinese. doi: 10.3969/j.issn.1008-8849.2016.05.002

**Li 2016**

1. Li M, Li J, Liu Y, et al. 盐酸小檗碱对精神分裂症患者糖脂代谢的影响 [The influence of berberine hydrochloride on glucolipid metabolism of patients with schizophrenia]. Chinese Journal of Prevention and Control of Chronic Diseases 2016. Chinese. doi: 10.16386/j.cjpccd.issn.1004-6194.2016.04.007
2. Li M, Liu Y, Qiu Y, et al. The effect of berberine adjunctive treatment on glycolipid metabolism in patients with schizophrenia: A randomized, double-blind, placebo-controlled clinical trial. Psychiatry Res 2021;300:113899. doi: 10.1016/j.psychres.2021.113899

**Wang 2016**

1. Wang L, Peng LY, Wei GH, Ge H. 黄连素胶囊治疗轻度高脂血症疗效观察[Therapeutic Effects of Berberine Capsule on Patients with Mild Hyperlipidemia]. Chinese Journal of Integrated Traditional and Western Medicine 2016;36:681-684. Chinese. doi:10.7661/CJIM.2016.06.0681

**Zhang 2016**

1. Zhang J, Zhao Y, Jia Q, et al. 黄连素对利培酮治疗精神分裂症患者症状及糖脂代谢的影响 [Effect of berberine on symptoms and glycolipid metabolism in schizophrenia patients treated with risperidone]. Journal of Tianjin Medical University 2016;22(4): 338-340. Chinese. Available from: https://d.wanfangdata.com.cn/periodical/tianjykdxxb201604017

**Zhu 2016**

1. Zhu Q, Hu W, Dai C. 小檗碱联合炔雌醇醋酸环丙孕酮治疗肥胖型多囊卵巢综合征疗效观察 [Effects of berberine combined with ethinylestradiol cyproterone acetate in the treatment of obese women with polycystic ovary syndrome]. Chinese Journal of Primary Medicine and Pharmacy 2016;23(6):837-840. Chinese. doi: 10.3760/cma.j.issn.1008-6706.2016.06.010

**Rashidi 2018**

1. Rashidi H, Namjoyan F, Mehraban Z, et al. The effects of active ingredients of barberry root (berberine) on glycemic control and insulin resistance in type 2 diabetic patients. Jundishapur Journal of Natural Pharmaceutical Products 2018;13(1). doi: 10.5812/jjnpp.64180

**Zhang 2020**

1. Zhang Y, Gu Y, Ren H, et al. Gut microbiome-related effects of berberine and probiotics on type 2 diabetes (the PREMOTE study). Nat Commun 2020;11(1):5015. doi: 10.1038/s41467-020-18414-8

**Hu 2021**

1. Hu J, Hu t, Wang j, et al. 盐酸小檗碱联合二甲双胍对2型糖尿病合并NAFLD患者体脂成分和肝脏脂肪含量的影响 [Effect of berberine combined with metformin on body fat distribution and liver fat content in type 2 diabetic patients with NAFLD]. Zhejiang Medicine 2021;43(21):2327-2331. Chinese. Available from: http://www.cnki.com.cn/Article/CJFDTOTAL-ZJYE202121013.htm

**Ming 2021**

1. Ming J, Xu S, Liu C, et al. Effectiveness and safety of bifidobacteria and berberine in people with hyperglycemia: study protocol for a randomized controlled trial. Trials 2018;19(1):72. doi: 10.1186/s13063-018-2438-5
2. Ming J, Yu X, Xu X, et al. Effectiveness and safety of *Bifidobacterium* and berberine in human hyperglycemia and their regulatory effect on the gut microbiota: a multi-center, double-blind, randomized, parallel-controlled study. Genome Med 2021;13(1):125. doi: 10.1186/s13073-021-00942-7

**Zhao 2021**

1. Zhao JV, Yeung WF, Chan YH, et al. Effect of berberine on cardiovascular disease risk factors: a mechanistic randomized controlled trial. Nutrients 2021;13(8). doi: 10.3390/nu13082550

### Supplementary Figures

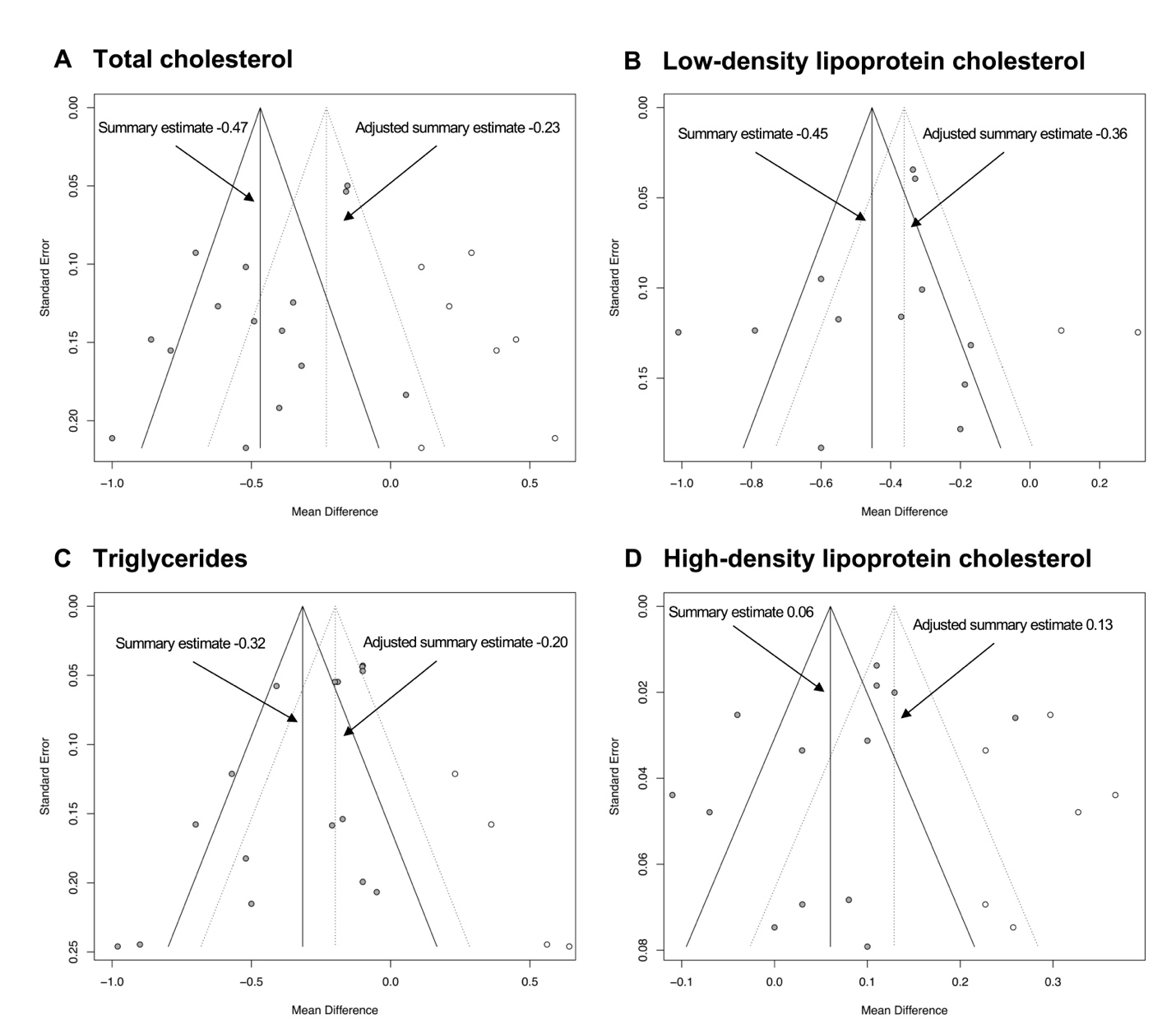
**Supplementary Figure 1**: Trim and fill funnel plots for total cholesterol (A), LDL cholesterol (B), HDL cholesterol (C) and triglycerides (D). Unfilled circles indicate the “filled” studies and the dashed vertical line shows the adjusted estimated mean difference.

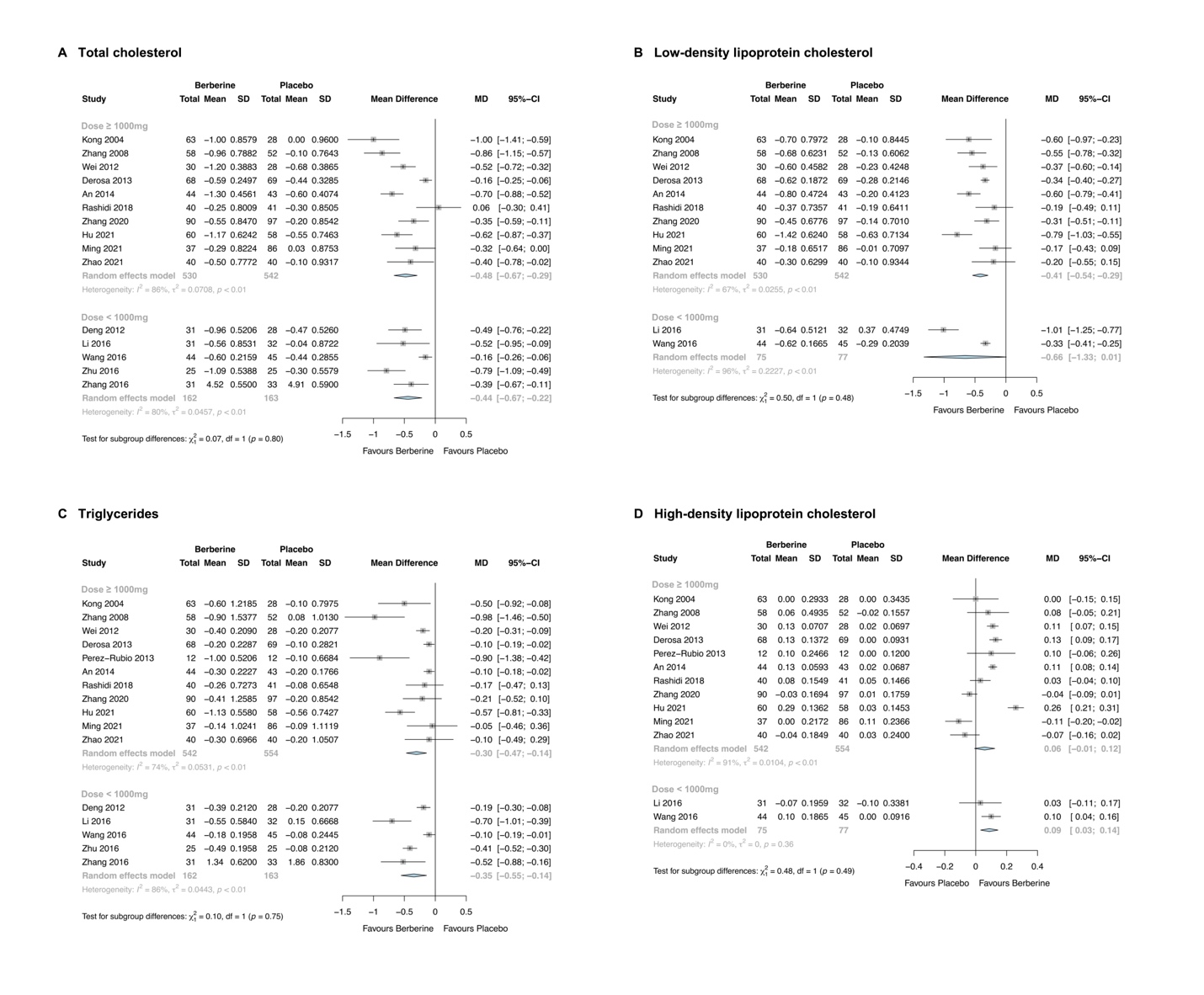
**Supplementary Figure 2**: Forest plot stratified by daily dose of berberine for total cholesterol (A), low-density lipoprotein cholesterol (B), triglycerides (C) and high-density lipoprotein cholesterol (D).

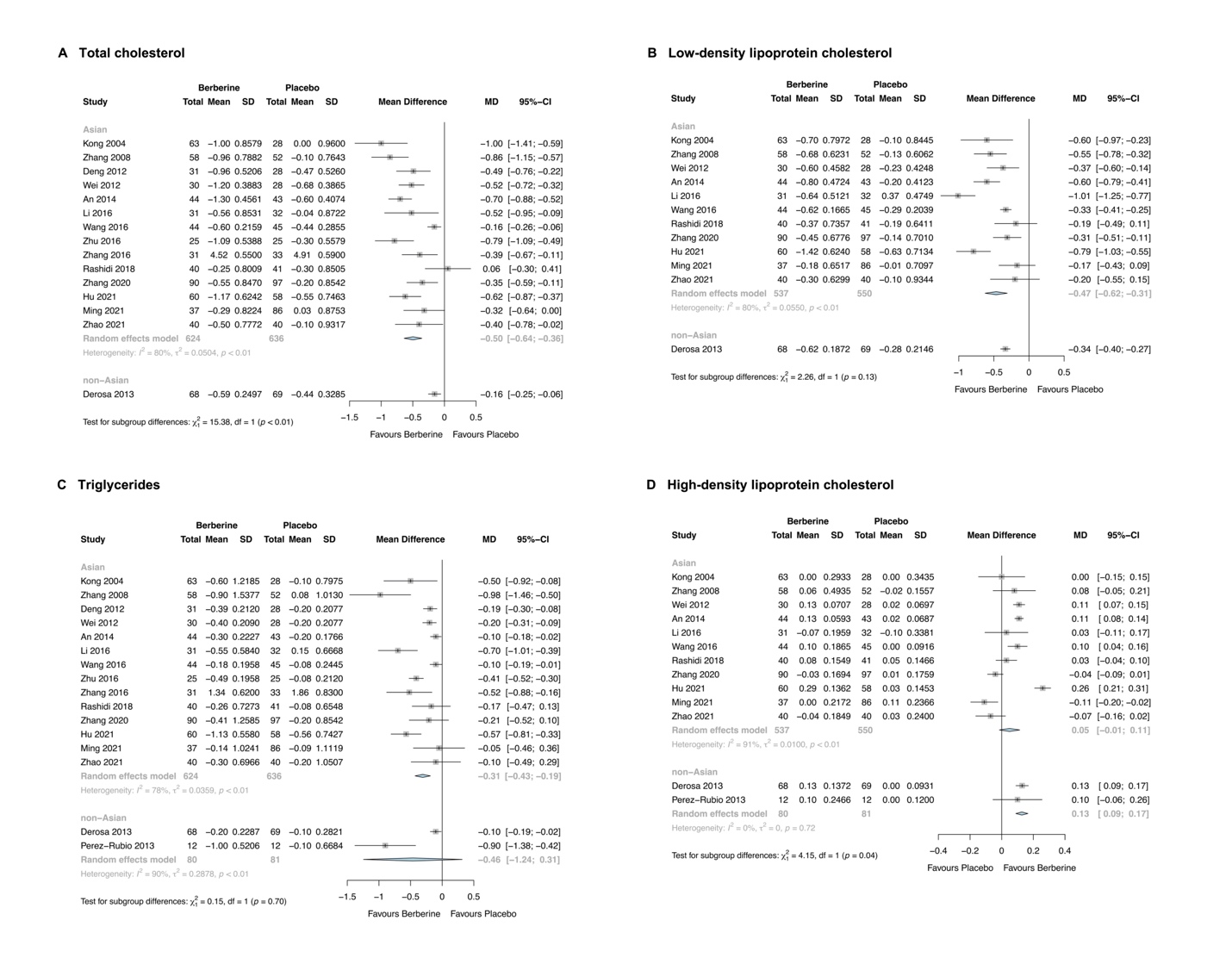
**Supplementary Figure 3:** Forest plot stratified by ethnicity for total cholesterol (A), low-density lipoprotein cholesterol (B), triglycerides (C) and high-density lipoprotein cholesterol (D).
