## Supplementary File 3 for "Overall and sex-specific effect of berberine for dyslipidemia: systematic review and meta-analysis of placebo-controlled trials"

### Preview

#### General information

Study ID

Last name and year

Year

Year of main publication

Journal Name

Name of journal for main publication

Title

Title of paper / abstract / report that data are extracted from

Lead author name

Last name, First name

Corresponding author name and email

Country in which the study was conducted

1. ○ China, mainland
2. ○ Hong Kong
3. ○ Italy
4. ○ Iran
5. ○ Taiwan
6. ○ United States
7. ○ Mexico
8. ○ Multiple countries
9. ○ Other

Study funding source

1. ☐ Government
2. ☐ Industry
3. ☐ Institutional
4. ☐ Not for profit

5. None
6. Other

Possible conflicts of interest for study authors

Clinical Trial Registration

Registry name and number

#### Study Design and Methods

#### Methods

Aim of study

Study design

1. Randomised controlled trial
2. Other

Start date

DD-MM-YYYY

End date

DD-MM-YYYY

Blinding (masking)

1. Unblinded
2. Single (participants)
3. Single (investigators)
4. Double (participants and investigators)
5. Triple (participants, investigators and analysts)
6. Other

Number of arms

1. Two (parallel)
2. Two (parallel with washout period)
3. Two (cross-over)

4. Three
5. Four
6. Other

Statistical model for analysis

#### Intervention and comparator

Intervention and Comparison

Meals: Yes or No

If not described: Not reported

Dosage form could be tablets, capsules, or other oral form.

| | Description | Dose | Frequency | Meals | Dosage form |
|---|---|---|---|---|---|
| **Berberine** | | | | | |
| **Placebo** | | | | | |

Treatment Duration

In weeks or months, preferably document number of weeks if reported.

Notes on intervention and comparator

Any additional interventions or procedures applied to one or both groups

#### Participants

Study population description

Total number of participants randomized

Number of participants in each arm

Excluded includes lost to follow-up and other reasons.

| | Allocated | Excluded | Analyzed |
|---|---|---|---|
| **Berberine** | | | |
| **Placebo** | | | |

Comments on participant flow

Baseline Population Characteristics

Extract % unless otherwise indicated.

HgA1c unit:

1 - %

2 - mmol/mol

| | Berberine | Placebo | Overall |
|---|---|---|---|
| **Female** | | | |
| **Male** | | | |
| **Ethnicity, Chinese** | | | |
| **Ethnicity, White/European** | | | |
| **Ethnicity, Other** | | | |
| **Age (mean/median)** | | | |
| **Current smoker** | | | |
| **Non-smoker (%)** | | | |
| **Diabetes mellitus (%)** | | | |
| **Hypertension (%)** | | | |
| **Coronary disease (%)** | | | |
| **Stroke (%)** | | | |
| **BMI (mean/median)** | | | |
| **HgA1c (mean/median)** | | | |
| **HgA1c, unit** | | | |

### Efficacy Outcomes

Measurement time and units

Outcome reported

0 - No

1 - Yes

Measure of central tendency

1 - Mean

2 - Median

Measure of distribution

1 - Standard deviation

2 - Standard error

3 - Interquartile range

4 - Range

Units

1 - mmol/L

2 - mg/dL

Time points for follow-up (extract in weeks):

- 1 month (4 weeks)

- 2 months (8 weeks)

- 3 months (12 weeks)

- 6 months (24 weeks) etc

| | Outcome reported | Measure of central tendency | Measure of distribution | Units | Time point 1 (t1) | Time point 2 (t2) | Time point 3 (t3) |
|---|---|---|---|---|---|---|---|
| **Total Cholesterol (TC)** | | | | | | | |
| **LDL cholesterol** | | | | | | | |
| **Triglycerides (TG)** | | | | | | | |
| **HDL cholesterol** | | | | | | | |
| **non-HDL cholesterol** | | | | | | | |
| **ApoB** | | | | | | | |

Efficacy outcomes

Change value (compares changes between two groups):

Extract the value of the difference between groups at the time point

Change spread, extract confidence interval: e.g. -0.70, -0.08.

| | Baseline central tendency Berberine | Baseline distribution Berberine | Baseline central tendency Placebo | Baseline distribution Placebo | Central tendency t1 Berberine | Distribution t1 Berberine | Central tendency t1 Placebo | Distribution t1 Placebo | Change, t1 | Change spread, t1 | Change p value, t1 | Central tendency t2 Berberine | Distribution t2 Berberine | Central tendency t2 Placebo | Distribution t2 Placebo | Change, t2 | Change spread, t2 | Change p value, t2 | C te Be |
|---|---|---|---|---|---|---|---|---|---|---|---|---|---|---|---|---|---|---|---|
| **Total cholesterol** | | | | | | | | | | | | | | | | | | | |
| **LDL cholesterol** | | | | | | | | | | | | | | | | | | | |

| | Baseline central tendency Berberine | Baseline distribution Berberine | Baseline central tendency Placebo | Baseline distribution Placebo | Central tendency t1 Berberine | Distribution t1 Berberine | Central tendency t1 Placebo | Distribution t1 Placebo | Change, t1 | Change spread, t1 | Change p value, t1 | Central tendency t2 Berberine | Distribution t2 Berberine | Central tendency t2 Placebo | Distribution t2 Placebo | Change, t2 | Change spread, t2 | Change p value, t2 | [illegible] |
|---|---|---|---|---|---|---|---|---|---|---|---|---|---|---|---|---|---|---|---|
| **Triglycerides** | | | | | | | | | | | | | | | | | | | |
| **HDL cholesterol** | | | | | | | | | | | | | | | | | | | |
| **non-HDL cholesterol** | | | | | | | | | | | | | | | | | | | |
| **ApoB** | | | | | | | | | | | | | | | | | | | |

#### Safety outcomes

Adverse events

If reported, extract both the number of adverse events (AE) and the number of participants/patients with the specific adverse event.

| | Number of events Berberine | Number of participants Berberine | Number of events Placebo | Number of participants Placebo |
|---|---|---|---|---|
| **Any adverse event** | | | | |
| **Drug-related adverse event** | | | | |
| **AE leading to discontinuation** | | | | |
| **Serious adverse event** | | | | |
| **Diarrhea** | | | | |
| **Constipation** | | | | |
| **Muscle pain** | | | | |
| **Rash** | | | | |
| **Any gastrointestinal AE** | | | | |
| **Nausea or vomiting** | | | | |
| **Nausea** | | | | |
| **Vomiting** | | | | |
| **Abdominal discomfort or GI pain** | | | | |
| **Flatulence** | | | | |
| **Hepatotoxicity/liver injury** | | | | |
| **Myopathy** | | | | |
| **Headache** | | | | |
| **Hypoglycemia** | | | | |
| **Burping or belching** | | | | |
| **Hyperglycemia** | | | | |
| **Weight gain** | | | | |

Notes on outcome reporting

Comment on scale, measures of central tendency, dispersion etc.
